# Prevalence of Macroprolactin in Hyperprolactinemic patients receiving Anti-psychotics

**DOI:** 10.1101/2021.05.11.21256928

**Authors:** Loai Ali Al Mortada Al Wasify, Shaikha Said Al Maamary, Mohammed Nasser Rashid Al Tobi

## Abstract

**Background:** Prolactin (PRL) hormone circulates in the blood in three forms, mono-prolactin which constitutes 85 % of prolactin in healthy and hyperprolactinemia conditions, a dimeric prolactin (big PRL) and polymeric PRL (big-big prolactin or Macroprolactin). Macroprolactin in normal conditions is not exceeding 2% of the total serum prolactin and had no biological activity. In some cases, of hyperprolactinemia the dominant form becomes MaPRL and exceeding the 2% percentage of total leading to misdiagnosis of hyperprolactinemia and un-necessary radiological investigations. The aim of this study is to detect the prevalence of MaPRL in Psychiatric patients with hyperprolactinemia due to anti-psychotic medications at Al Masarra hospital.

**Material and methods:** The study was conducted on 190 samples from patients with high prolactin in Al Masarra Hospital either inpatients or out-patient clinics either male or non-lactating not pregnant female. The measurement of Prolactin level was measured by the automated analyser COBAS e411, Roche Diagnostic. Macroprolactin was precipitated by using Polyethylene glycol (PEG).

**Results:** Prevalence of Macroprolactin was 10.5 % of hyperprolactinemic patients receiving antipsychotics. There was a statistically significant difference in gender between the symptomatic and asymptomatic group. There was no significant difference in medications used between the symptomatic and asymptomatic group and there was a statistically significant difference in total Prolactin & Macroprolactin between males and females.

**Conclusions:** Investigation for Macroprolactin should be done in every hyperprolactinemia patient who is receiving antipsychotics more especially the asymptomatic cases to avoid unnecessary radiological imaging and treatment.

## Introduction

PRL is produced primarily by pituitary lactotrophs and is tonically inhibited by hypothalamic dopamine. PRL is a 199–amino acid polypeptide containing three intramolecular disulfide bonds (1).

Prolactin contributes to hundreds of physiologic functions, but the two primary responsibilities are milk production and development of mammary glands within breast tissues (2).

PRL facilitates the maturation of T cells via IL-2 receptor expression, impairs B cell tolerance to self-antigens through the anti-apoptotic effect, develops antigen-presenting cells, and enhances immunoglobulin production (3).

Prolactin circulates in the blood in different forms; monomeric PRL with molecular weight 23 kDa (referred to as little PRL),, dimeric PRL with molecular weight 48 to 56 kDa (big PRL) which is biologically inactive, and polymeric form of PRL with a molecular weight > 150 kDa (big-big PRL) which is also called a macroprolactin (MaPRL)

In the normal sera, monomeric PRL accounts for 80–95% of the total PRL, dimeric PRL makes up <10%, and macroprolactin accounts for a small amount of less than 1% of the total PRL (4).

The monomeric PRL represents the major circulatory isoform of the total PRL in cases with normoprolactinemia and true hyperprolactinemia. The biological and immunological activity of PRL may be almost exclusively attributed to the monomeric form (5).

The increase in serum Prolactin concentration (hyperprolactinemia) is caused physiologically by pregnancy, lactation, stress & pain and pathologically by pituitary adenoma (secreting PRL), hypothyroidism, chest wall disease, hepatorenal disorders and drug induced (anti-dopaminergic drugs).

The most common drugs that causes hyperprolactinemia are the antipsychotic medicines like respiredone, haloperidol, and olanzapine (6).

However, 29% of hyperprolactinemia has been classified as idiopathic, because the causes are unknown. Anti-PRL autoantibody was found to be one of the major causes of idiopathic hyperprolactinemia (7).

In most cases, MaPRL is composed of immune complexes of PRL and anti-PRL auto-antibodies, and 87% of MaPRL was PRL-IgG complex and 67% of MaPRL was autoantibody-bound PRL. Anti-PRL autoantibody-bound PRL is a major form of PRL-IgG complex and PRL – IgG complex is a major form of macroprolactin (8).

It has been shown that MaPRL is biologically inactive because the large molecular size of this complex prevents its crossing through the capillary blood barrier and reaching target cells and immunoglobulin connecting with specific epitops of PRL molecule may reduce the binding of the hormone to its receptors (9, 10, 20).

So, MaPRL has longer renal clearance resulting in its accumulation in the serum. In addition to, the most available immunoassays used to measure PRL level do not distinguish MaPRL from monomeric PRL, which can lead to an incorrect diagnosis of hyperprolactinaemia and un-necessary imaging study and treatment.

If MaPRL has been found to predominate in human serum, it will be called macroprolactinemia. In many studies, it is found in 10 – 45% of hyperprolactinemic patients, depending on the immunoassay used in the laboratory (11, 12, 13, 14).

Although most of the studies reported that MaPRL is inactive, some studies reported that patients with a high concentration of MaPRL exhibit signs or symptoms of hyperprolactinemia such as galactorrhea, menstrual irregularities, or infertility, other studies reported that macroprolactinemia cannot be differentiated from hyper PRL based on clinical symptoms alone, because several of the signs and symptoms of hyperprolactinemia are non-specific, it is possible that the occurrence of symptoms with macroprolactinemia is coincidental (15, 16, 17).

It is important to identify the presence of MaPRL as the cause of hyperprolactinemia to avoid unnecessary radiological investigations and treatment with dopamine agonist medicines.

Macroprolactin is found to interfere with most commercially available immunoassays used for prolactin. As a result, false high prolactin values (apparent hyperprolactinemia) are obtained and these values depend on the assay method employed. So, the most available immunoassays used to measure PRL level do not distinguish MaPRL from monomeric PRL, which can lead to an incorrect diagnosis of hyperprolactinaemia and un-necessary imaging study and treatment (18).

For detection of MaPRL, Gel filtration chromatography (GFC) is the gold standard method for quantification of the three variants of PRL, but this method is costly, time consuming and labor intensive, so it is not used in routine screening for MaPRL (19).

Laboratories generally use polyethylene glycol (PEG) precipitation to differentiate macroprolactinemia from true hyperprolactinemia. This method is simple and inexpensive and has been extensively validated against GFC.

The aim of this study is to evaluate the prevalence of MaPRL in hyperprolactinemic patients in Al Masarra hospital who are receiving anti-psychotic medications by precipitating MaPRL with PEG.

## Materials & Methods

### Patients

The study was done on 190 samples from patients with high prolactin level and were receiving anti-psychotics either male or non-pregnant non-lactating female from outpatient clinics or in-patients wards of the Al Masarra hospital which is a tertiary psychiatric hospital in Sultanate of Oman from March 2020 to August 2020.

### Immunoassay

The measurement of Prolactin level was measured by the automated analyser COBAS e411, Roche Diagnostic.

By using the electrochemiluminescence immunoassay “ECLIA” based on the sandwich principle, using biotinylated monoclonal specific antibody interacted with streptavidin-coated micro particles and second antibody which is monoclonal antibody labelled with a ruthenium complex. After application of voltage, the chemiluminescent emissions were measured.

The samples were separated and stored at -70 C^°^ till processing.

The reference range was: 102 – 496 µIU/mL for non-pregnant women and 86 – 324 µIU/mL for men.

The inter-assay coefficient of variation (CV) was 2.2% and the intra-assay was 2.5%at level.

### PEG-Precipitation

Preparation of 25% solution of PEG 6000 was prepared by dissolving 25 grams of PEG 6000 in 60 ml of distilled water at room temperature and mixing with vortex, then fulfilling the volumes till 100 ml of solution. The prepared solution is stable for three months maximum period at 4C^°^.

In a separate glass tube, equal amount of serum sample and 25% of PEG were added. After thorough vortex mixing for a minute and stabilization for 30 minutes, the solution was centrifuged at 9500 g for 10 minutes.

The supernatant portion of the sample was separated and prolactin level was measured in it.

The prolactin level in the treated sample was multiplied by two to correct for the dilution with PEG.

We followed the same protocol of Ana Maia Silva et al; 2014 in preparation of PEG solution and samples (20).

### Measuring Free-PRL

The free prolactin is the prolactin level which will be recovered after precipitation of MaPRL by PEG.

The macroprolactin level will be determined using the following formula:

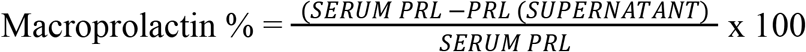

#### Interpretation

Free prolactin recovering less than 40% = significant presence of MaPRL. Free PRL recovering that will exceed 50% = Monomeric PRL predominance. Free PRL recovering between 40% -50% = intermediate or at the grey zone (21, 22, 23).

### Statistical Analysis

SPSS version 20 was used for data analysis. The level of statistically significant differences was set at *p* < 0.05.

## Results

The study showed that of 190 patients the mean age was 33.2 ± 9.2 years with the age range 16 – 62 years. There were 149 females (78.4 %) and 41 males (21.6%). 169 (88.9%) were Omani and 21 (11.1%) were non-Omani.

The study found that 75.8% of hyperprolactinaemia patients were asymptomatic and 24.2% were symptomatic.

56.6% of patients received risperidone, (29.3%) of patients received haloperidol, (9.4%) of patients received flupentixol decanoate and (4.7%) of patients received olanzapine.

We found that (10.5%) of hyperprolactinaemic patients had macroprolactin predominance with recovery rate (RR) between 25.6% -39.8% and (3.2%) of cases were borderline with RR between 44.3% - 50%, while (86.3%) of cases had free PRL with RR between 55 % - 98.9%.

25% of patients with MaPRL predominance were males and 75% were females. There was a statistically significant difference in gender between the symptomatic and asymptomatic group as males represented (28.5%) of the studied sample in the asymptomatic group and (0.0%) in the symptomatic group while females were (71.5%) in asymptomatic group in comparison to (100.0%) in symptomatic group (*p*=0.00).

There was a statistically significant difference in nationality between the symptomatic and asymptomatic group as Omani patients represented (92.4%) of the studied sample in asymptomatic group and (78.3%) in symptomatic group while non-Omani were (7.6%) in asymptomatic group in comparison to (21.7%) in symptomatic group (*p*=0.00).

There was a statistically significant difference in interpretation of hyperprolactinemia between the symptomatic and asymptomatic group as patients with free prolactin represented (86.1%) of the studied sample in the asymptomatic group and (87.0%) in the symptomatic group.

Meanwhile patients with macroprolactin were (13.2%) in the asymptomatic group in comparison to (2.2%) in the symptomatic group while patients with borderline level were (0.7%) in the asymptomatic group in comparison to (10.9%) in the symptomatic group (*p*=0.00).

There was a statistically significant difference between total prolactin and post -PEG PRL in which the mean of total PRL was 2413.6 ± 1486.0 and the mean of post-PEG PRL was 1759.37 ± 1260.5 (*p* = 0.000) which was in accordance of the result of Nedjeljka Ruljancic et al 2021 (24).

There was a statistically significant difference in total Prolactin between male and female in which the mean for female (2689.7 ± 1552.8) were significantly higher than in males (1410.4 ± 443.9) (*p*= 0.000).

There was also a statistically significant difference in macroprolactin level between male and female in which the mean for female (721.7 ± 495.3) were significantly higher than in males (408.9± 290.8) (*p* = 0.000), which was in line with the study of Young-Min Park et al 2016 (25).

There was a statistically significant difference in type of prolactin between the symptomatic and asymptomatic group as total prolactin mean was 2208.3 of the studied sample in the asymptomatic group and was 3056.1 in the symptomatic group while the mean of corrected prolactin were 1589.2 in the asymptomatic group in comparison to 2292 in the symptomatic group (*t-test* =0.001).

There was no significant difference in the mean of MaPRL between the asymptomatic and the symptomatic group (*t-test* =0.007).

The treatment of samples with PEG produced a reduction in PRL level in all cases with a mean reduction of 28.3%.

There was no significant difference in medications used between the symptomatic and asymptomatic group.

Risperidone represented (81.3%) of the studied sample in the asymptomatic group and (18.7%) in the symptomatic group while haloperidol represented (64.3%) in the asymptomatic group in comparison to (35.7%) in the symptomatic group.

Flupenthixol Decanoate represented (72.2%) in the asymptomatic group in comparison to (27.8%) in the symptomatic group and olanzapine represented (88.9%) in the asymptomatic group in comparison to (11.1%) in the symptomatic group (*p*=0.079) which was on the contrary of the study of Young-Min Park et al 2016 (25).

The mean of total PRL & MaPRL was higher in patients who were receiving haloperidol more than the other medications. (Table I) (Figure I).

**Table (I).** Mean of Total PRL & MaPRL of Olanzapine, Haloperidol, and Risperidone & Flupenthixol Decanoate.

|  | Olanzapine (n=9) |  |  | Haloperidol (n=56) |  |  | Risperidone (n=107) |  |  | Flupenthixol Decanoate (n=18) |  |  |
| --- | --- | --- | --- | --- | --- | --- | --- | --- | --- | --- | --- | --- |
|  | YES<br>Mean<br>± SD | NO<br>Mean<br>± SD | Sig. | YES<br>Mean ±<br>SD | NO<br>Mean ±<br>SD | Sig. | YES<br>Mean ±<br>SD | NO<br>Mean ±<br>SD | Sig. | YES<br>Mean<br>± SD | NO<br>Mean<br>± SD | Sig. |
| Total<br>PRL | 1710.7<br>± 893.7 | 2428.4<br>± 1482.0 | $t = -1.438$<br>$p = 0.152$ | 2560.9<br>± 1691.1 | 2331.4<br>± 1358.9 | $t = 1.021$<br>$p = 0.309$ | 2388.3 ±<br>1397.5 | 2446.2<br>± 1601 | $t = 0.265$<br>$p = 0.791$ | 1829.4<br>± 880.5 | 2478.4<br>± 1530.5 | $t = -1.765$<br>$p = 0.079$ |
| MaPRL | 679.7<br>± 664.9 | 650.9<br>± 467.7 | $t = -0.177$<br>$p = 0.860$ | 700.8<br>± 467.5 | 628.2<br>± 480.6 | $t = -1.007$<br>$p = 0.315$ | 628.2<br>± 474.0 | 687.7<br>± 479.4 | $t = -0.853$<br>$p = 0.395$ | 567.0 ±<br>316.5 | 665.8 ±<br>491.7 | $t = -0.833$<br>$p = 0.406$ |

|  | Olanzapine (n=9) |  |  | Haloperidol (n=56) |  |  | Risperidone (n=107) |  |  | Flupenthixol Decanoate (n=18) |  |  |
| --- | --- | --- | --- | --- | --- | --- | --- | --- | --- | --- | --- | --- |
|  | YES<br>Mean<br>± SD | NO<br>Mean<br>± SD | Sig. | YES<br>Mean ±<br>SD | NO<br>Mean ±<br>SD | Sig. | YES<br>Mean ±<br>SD | NO<br>Mean ±<br>SD | Sig. | YES<br>Mean<br>± SD | NO<br>Mean<br>± SD | Sig. |
| Total<br>PRL | 1710.7<br>± 893.7 | 2428.4<br>± 1482.0 | T= -<br>1.438<br>P=<br>0.152 | 2560.9<br>±1691.1 | 2331.4<br>± 1358.9 | T= 1.021<br>P=0.309 | 2388.3<br>± 1397.5 | 2446.2<br>± 1601 | T=0.265<br>P=0.791 | 1829.4<br>± 880.5 | 2478.4<br>± 1530.5 | T= -<br>1.765<br>P=0.079 |
| MaPRL | 679.7<br>± 664.9 | 650.9<br>± 467.7 | T= -<br>0.177<br>P=<br>0.860 | 700.8<br>± 467.5 | 628.2<br>± 480.6 | T=-1.007<br>P=0.315 | 628.2<br>± 474.0 | 687.7<br>± 479.4 | T= -<br>0.853<br>P=0.395 | 567.0<br>± 316.5 | 665.8<br>± 491.7 | T= -<br>0.833<br>P= 0.406 |

**Figure (I).**
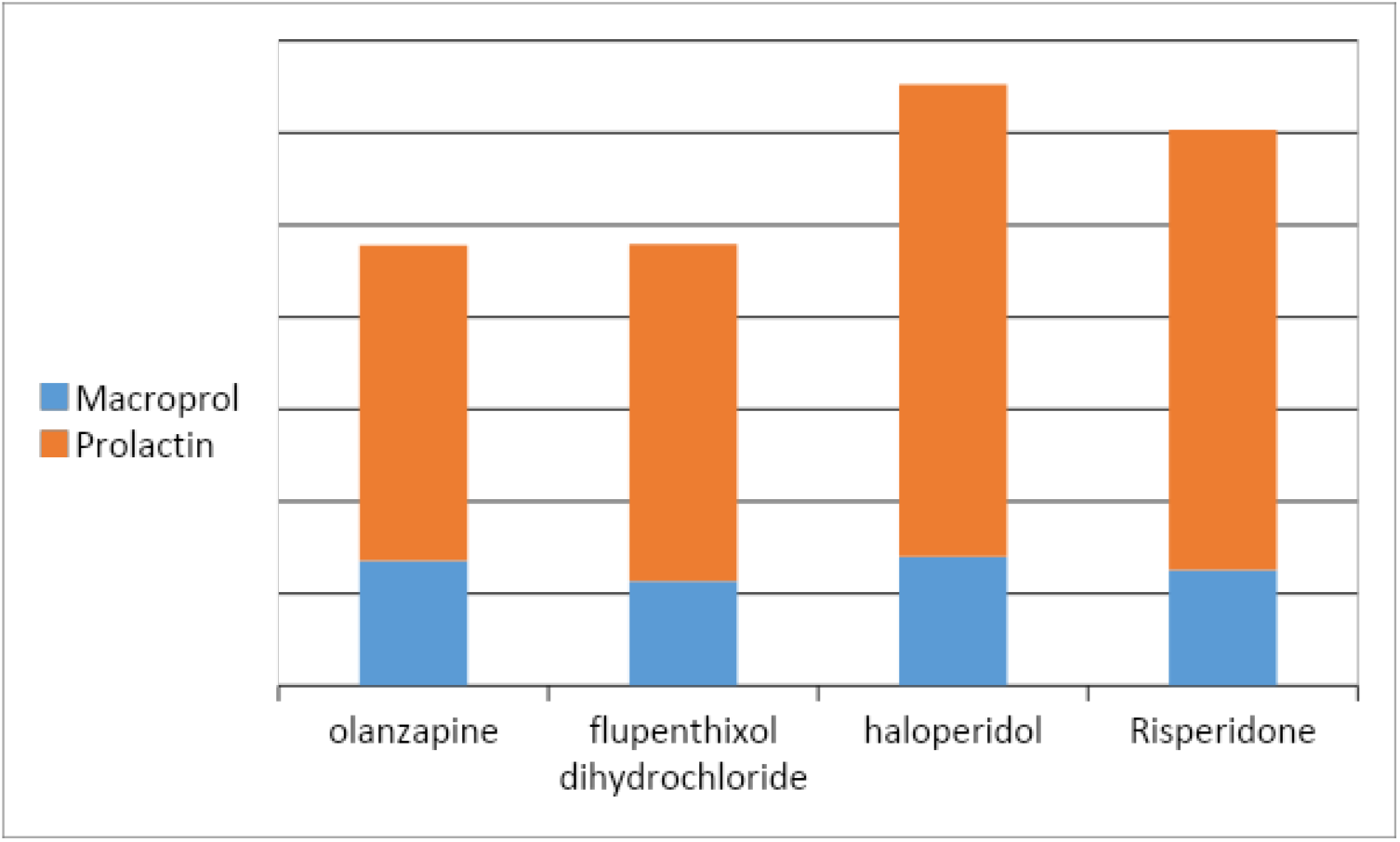
the mean of PRL & MaPRL in Olanzapine, Flupenthixol Decanoate, Haloperidol & Risperidone

In our study there was a positive correlation between total prolactin and macroprolactin levels (*r* = 0.59, *p* < 0.01). (Figure II).

**Figure (II).**
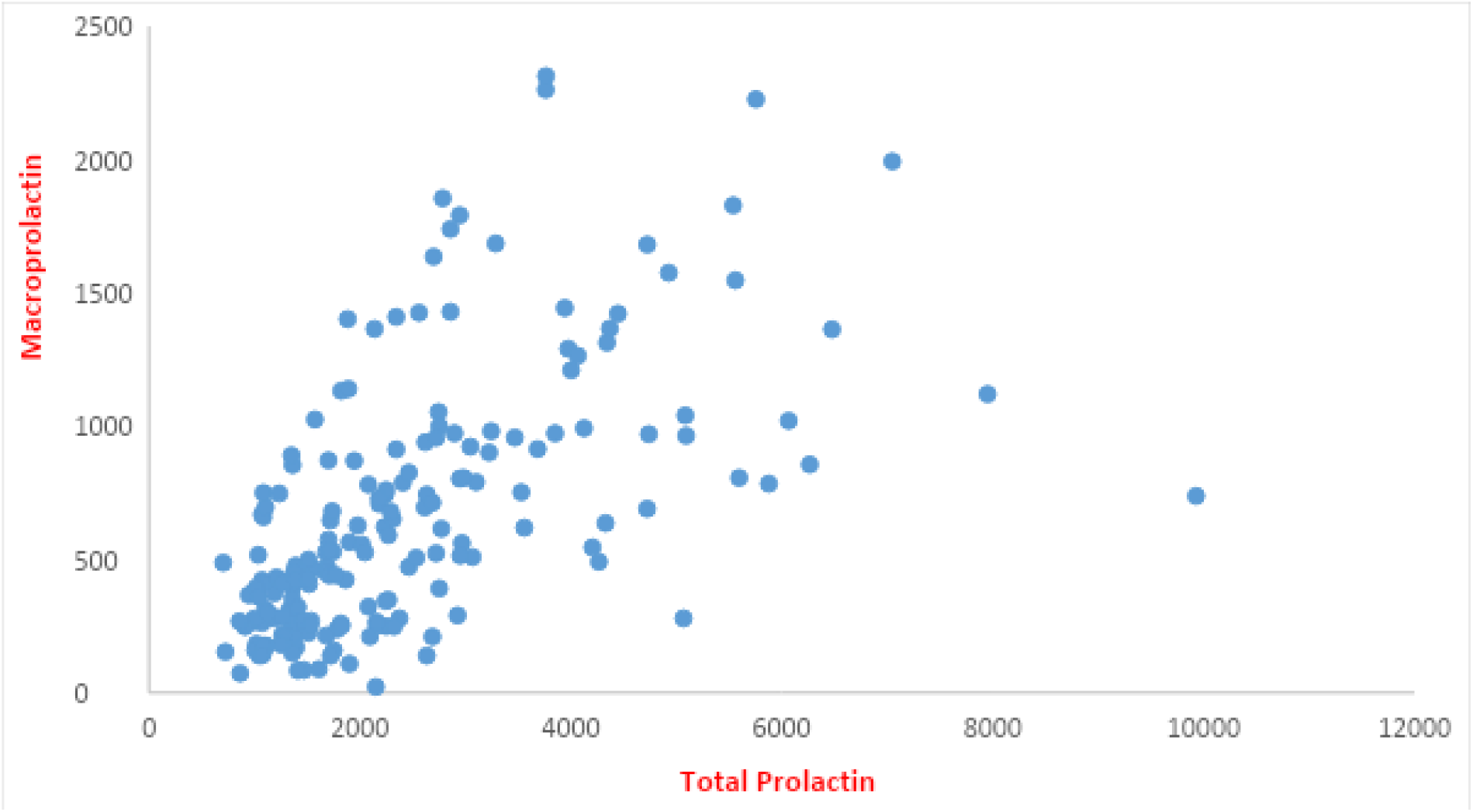
Correlation between total PRL & MaPRL.

## Discussion

Macroprolactin was described for the first time by Wittaker et al in 1981, when he reported a case with hyperprolactinemia without the common symptoms of amenorrhea, galactorrhea and infertility. It was found that the high molecular weight MaPRL accounted for the majority of PRL in this patient by using GFC. It is PRL-IgG immunocomplex in which endogenous IgG molecule is directed against epitopes on N- and C-terminal residues of monomeric PRL (26).

In the present study, treatment of the sera of patients with high prolactin with 25 % PEG 6000 was done leading to precipitation of macroprolactin and free prolactin level was measured in the supernatant. This method is the easiest and cheapest method for detecting MaPRL but it can precipitate monomeric PRL, also with MaPRL leads to reduction of post-PEG PRL values in all serum samples with a mean reduction of 28.3% in this study.

In our study we used the most common cut-off <40 % recovery rate to consider the case positive for macroprolactinemia. It has been reported that the 40% cutoff is 100% sensitive to confirm the presence of MaPRL, while using a 60% cutoff does not report significantly more patients with significant macroprolactinemia (26).

In our study, 10.5% were detected with macroprolactin predominance which is consistent with studies that have been documented in the United States which found that about 10% of hyperprolactinemic patients have MaPRL as a predominant form of PRL, however most of the studies conducted in Europe have shown that the predominance of MaPRL is usually above 20% (27, 28, 29, 30).

95 % of cases of MaPRL were asymptomatic and 5 % were complaining of amenorrhea. Although MaPRL is biologically inactive but the symptoms can be a coincidence or an intermittent dissociation of macroprolactin from IgG molecule could happen, causing the symptoms as suggested by Hattori N et al in 1997 and M Kasum et al. (29,5).

Suliman AM et al also explained that although Big-Big PRL may be the predominant molecular form in MaPRL, monomeric PRL may simultaneously be found in excess. Therefore, MaPRL could be associated with monomeric hyperprolactinemia which leads to the development of hyperprolactinemic symptoms (31).

In our study prolactin levels and macroprolactin levels were significantly higher in females than males like previous results of Johnsen et al 2008 & Young-Min Park et al 2016 (31-32).

The antipsychotics induced hyperprolactinemia is related to differential D2 receptor affinity, 5-hydroxytryptamine receptor affinity and blood-brain disposition of antipsychotics (33 - 34).

Risperidone is one of the antipsychotics that has poor blood-brain barrier penetration and high concentrations of risperidone exist in the pituitary and has a strong D2-blocking effect and a simultaneous effect on the 5-HT_2_ receptors (35).

In our study risperidone appeared to increase prolactin in 56.3% of patients, while haloperidol increased PRL in 29.5% of patients, however flupentixol decanoate increased PRL in 9.5% of patients, meanwhile Olanzapine increased PRL in 4.7 % of patients.

In our study there was no significant difference in medications used between the symptomatic and asymptomatic groups.

The mean prolactin and macroprolactin levels were higher in patients who was receiving haloperidol than the other medicines but was no statistically significant difference between them.

Beda-Maluga et al reported that most patients with a PRL concentration higher than 100 ng/ml (2127 uIU/mL) had true hyperprolactinemia, although significant macroprolactinemia might be present in some cases (36).

The American Association of Clinical Endocrinologists recommends using a 25–150 ng/mL (532 - 3191 uIU /mL) interval to search for MaPRL, and the Endocrine Society suggests testing every asymptomatic hyperprolactinemic patient for MaPRL (37).

## Conclusion

Macroprolactin predominance in hyperprolactinemic patients receiving antipsychotics was 10.5 % which is in line with many literatures. Haloperidol was associated with increased total and macroprolactin levels more than other drugs. So, we recommend an investigation for macroprolactin predominance in every hyperprolactinemic patient, especially the asymptomatic cases to avoid unnecessary radiological imaging and treatment.

## Ethical considerations

The authors declare that they got approval from the hospital administration and DGMS to access the electronic files of the patients and collect the medical history and data.

The authors declare that no patient data appear in this article.

In this research there was no direct contact with patients, so there was no need for their consent.

## Supporting information

Ethical approval

## Data Availability

The authors declare that all data of the study are available upon request.

## Acknowledgment

The authors would like to thank Dr. Sameh Yousry Bondok for his valuable help and support in this study.

## Abbreviations

PRL: Prolactin
MaPRL: Macroprolactin
PEG: Polyethylene glycol
IgG: Immunoglobulin G
RR: Recovery rate
GFC: Gel Filtration Chromatography

## References

1. Melmed S, Koenig RJ, Auchus RJ, Rosen CJ; Allison B. Goldfine. Williams Textbook of Endocrinology. Philadelphia: Saunders; 2019. Physiology and Disorders of Pituitary Hormone Axes; p. 190.

2. Al-Chalabi M, Bass AN, Alsalman I. Physiology, Prolactin. In: StatPearls. Treasure Island (FL): StatPearls Publishing; July 10, 2020.

3. S. Shelly, M. Boaz, and H. Orbach, “Prolactin and autoimmunity,” Autoimmunity Reviews, vol. 11, no. 6-7, pp. A465–A470, 2012.

4. Che Soh, N.A.A.; Yaacob, N.M.; Omar, J.; Mohammed Jelani, A.; Shafii, N.; Tuan Ismail, T.S.; Wan Azman, W.N.; Ghazali, A.K. Global Prevalence of Macroprolactinemia among Patients with Hyperprolactinemia: A Systematic Review and Meta-Analysis. Int. J. Environ. Res. Public Health 2020, 17, 8199.

5. M Kasum, S. Oreskovic, E Čehić,, M Šunj, A. Lila, E. Ejubovic.Laboratory and clinical significance of macroprolactinemia in women with hyperprolactinemia. Taiwanese Journal of Obstetrics and Gynecology 2017, Volume 56, Issue 6, 719–724.

6. Majumdar A, Mangal NS. Hyperprolactinemia. J Hum Reprod Sci. 2013; 6(3):168-175. doi:10.4103/0974-1208.121400

7. Shimatisu A., Hattori N, “Macroprolactinemia: Diagnostic, Clinical, and Pathogenic Significance”, Journal of Immunology Research, Vol.2012, Article ID 167132, 7 pages.

8. Hattori, N., Ishihara, T., Saiki, Y., & Shimatsu, A. (2010). Macroprolactinaemia in patients with hyperprolactinaemia: composition of macroprolactin and stability during long-term follow-up. Clinical endocrinology, 73(6), 792–797.

9. Hattori N. (2003). Macroprolactinemia: a new cause of hyperprolactinemia. Journal of pharmacological sciences, 92(3), 171–177.

10. Hattori N, Nakayama Y, Kitagawa K et al. Development of anti-PRL (prolactin) autoantibodies by homologous PRL in rats: a model for macroprolactinemia. Endocrinology 2007; 148: 2465–2470.

11. Ceratto, K.M., Zitta, M.M., Avalos, P., Gomez, M.H., Avendano, C., Sanchez Sarmiento Cesar. (2016). Prevalence of macroprolactin (MPRL) in infertile female patients: frequent misdiagnosis and mismanagement of hyperprolactinemia. Fertility and Sterility 2016, 106.e246.10.1016/j.fertnstert.2016.07.710.

12. Thirunavakkarasu, K., Dutta, P., Sridhar, S., Dhaliwal, L., Prashad, G. R., Gainder, S., Sachdeva, N., & Bhansali, A. (2013). Macroprolactinemia in hyperprolactinemic infertile women. Endocrine, 44(3), 750–755.

13. Samson, S. L., Hamrahian, A. H., Ezzat, S., AACE Neuroendocrine and Pituitary Scientific Committee, & American College of Endocrinology (ACE) (2015). AMERICAN ASSOCIATION OF CLINICAL ENDOCRINOLOGISTS, AMERICAN COLLEGE OF ENDOCRINOLOGY DISEASE STATE CLINICAL REVIEW: CLINICAL RELEVANCE OF MACROPROLACTIN IN THE ABSENCE OR PRESENCE OF TRUE HYPERPROLACTINEMIA. Endocrine practice: official journal of the American College of Endocrinology and the American Association of Clinical Endocrinologists, 21(12), 1427–1435.

14. Kalsi AK, Halder A, Jain M, Chaturvedi PK, Sharma JB. Prevalence and reproductive manifestations of macroprolactinemia. Endocrine. 2019 Feb; 63(2):332–340. DOI: 10.1007/s12020-018-1770-6.

15. Vilar, L., Abucham, J., Albuquerque, J. L., Araujo, L. A., Azevedo, M. F., Boguszewski, C. L., Casulari, L. A., Cunha Neto, M., Czepielewski, M. A., Duarte, F., Faria, M., Gadelha, M. R., Garmes, H. M., Glezer, A., Gurgel, M. H., Jallad, R. S., Martins, M., Miranda, P., Montenegro, R. M., Musolino, N., Bronstein, M. D. (2018). Controversial issues in the management of hyperprolactinemia and prolactinomas - An overview by the Neuroendocrinology Department of the Brazilian Society of Endocrinology and Metabolism. Archives of endocrinology and metabolism, 62(2), 236–263

16. Smith, T. P., Kavanagh, L., Healy, M. L., & McKenna, T. J. (2007). Technology insight: measuring prolactin in clinical samples. Nature clinical practice. Endocrinology & metabolism, 3(3), 279–289.

17. Gibney, J., Smith, T. P., & McKenna, T. J. (2005). The impact on clinical practice of routine screening for macroprolactin. The Journal of clinical endocrinology and metabolism, 90(7), 3927–3932.

18. Haddad RA, Giacherio D, Barkan AL. Interpretation of common endocrine laboratory tests: technical pitfalls, their mechanisms and practical considerations. Clin Diabetes Endocrinol. 2019; 5:12. Published 2019 Jul 24. doi: 10.1186/s40842-019-0086-7.

19. Fahie-Wilson, M., & Smith, T. P. (2013). Determination of prolactin: the macroprolactin problem. Best practice & research. Clinical endocrinology & metabolism, 27(5), 725–742.

20. Ana Maia Silva, Paula Martins da Costa, Ana Pacheo, Jose Carlos Oliveria & Claudia Freitas. Assessment of macroprolactinemia by polyethylene glycol precipitation method: Rev Port Endocrinol Diabetes Metab2014; 9(1):25–28.

21. Guclu, M., Cander, S., Kiyici, S. et al. Serum macroprolactin levels in pregnancy and association with thyroid autoimmunity. BMC Endocr Disord 15, 31 (2015).

22. Gilson G, Schmit P, Thix J, Hoffman J, Humbel R. Prolactin results for samples containing macroprolactin are method and sample dependent. Clin Chem 2001; 47: 331–333.

23. Che Soh Naa, Yaacob NM, Omar J, Mohammed Jelani A, Shafii N, Tuan Ismail TS, Wan Azman WN, Ghazali AK. Global Prevalence of Macroprolactinemia among Patients with Hyperprolactinemia: A Systematic Review and Meta-Analysis. International Journal of Environmental Research and Public Health. 2020; 17(21):8199. https://doi.org/10.3390/ijerph17218199

24. Nedjeljka Ruljancic, Ana Bakliza1, Sandra Vuk Pisk, Natko Geres, Katarina Matic, Ena Ivezic, Vladimir Grosic, Igor Filipcic; Antipsychotics-induced hyperprolactinemia and screening for macroprolactin, Biochem Med (Zagreb). 2021 Feb 15; 31(1): 010707.

25. Park, Y. M., Lee, S. H., Lee, B. H., Lee, K. Y., Lee, K. S., Kang, S. G., Lee, H. Y., & Kim, W. (2016). Prolactin and macroprolactin levels in psychiatric patients receiving atypical antipsychotics: A preliminary study. Psychiatry research, 239, 184–189.

26. Wittaker PG. Wilcox T, Lind T. Maintained fertility in a patient with hyperprolactinemia due to big – big prolactin. J Clin Endocrinol Metab. 1981; 53(4): 863–6.

27. Parlant-Pinet, L., Harthé, C., Roucher, F., Morel, Y., Borson-Chazot, F., Raverot, G., & Raverot, V. (2015). Macroprolactinaemia: a biological diagnostic strategy from the study of 222 patients. European journal of endocrinology, 172(6), 687–695.

28. Sapin R, Kertesz G. Macroprolactin detection by precipitation with Protein a —Sepharose: a rapid screening method compared with polyethylene glycol precipitation. Clin Chem 2003; 49: 502–504.

29. Hattori N, Inagaki C. Anti-prolactin (PRL) autoantibodies cause asymptomatic hyperprolactinemia: bioassay and clearance studies of PRL-immunoglobulin G complex. J Clin Endocrinol Metab. 1997 Sep; 82(9):3107–10. doi: 10.1210/jcem.82.9.4250. PMID: 9284753

30. Suliman AM, Smith TP, Gibney J, McKenna TJ. Frequent misdiagnosis and mismanagement of hyperprolactinemic patients before the introduction of macroprolactin screening: application of a new strict laboratory definition of macroprolactinemia. Clin Chem. 2003Sep; 49(9):1504-9.doi: 10.1373/49.9.1504. PMID: 12928232.

31. Kinon, B. J., Gilmore, J. A., Liu, H., & Halbreich, U. M. (2003). Hyperprolactinemia in response to antipsychotic drugs: characterization across comparative clinical trials. Psychoneuroendocrinology, 28 Suppl 2, 69–82.

32. Johnsen, E., Kroken, R. A., Abaza, M., Olberg, H., & Jørgensen, H. A. (2008). Antipsychotic-induced hyperprolactinemia: a cross-sectional survey. Journal of clinical psychopharmacology, 28(6), 686–690.

33. Fitzgerald, P., & Dinan, T. G. (2008). Prolactin and dopamine: what is the connection? A review article. Journal of psychopharmacology (Oxford, England), 22(2 Suppl), 12–19.

34. Kenshi T., Yurika Y., Hitoshi K., Mamoru T., Shingo T., Miwako K., Ichiro M., Hiroaki Y, Yoshito Z., Masaki I., Keisuke I., Akihiro T., Hiroaki A., Psychiatric Patients with Antipsychotic Drug-Induced Hyperprolactinemia and Menstruation Disorders, Biological and Pharmaceutical Bulletin, 2017, Volume 40, Issue 10, Pages 1775–1778, Released October 01, 2017.

35. Just MJ. The influence of atypical antipsychotic drugs on sexual function. Neuropsychiatr. Dis. Treat., 11, 1655–1661 (2015)

36. Beda-Maluga K, Pisarek H, Komorowski J, Swietoslawski J, Fuss Chmielewska J, Winczyk K. Evaluation of hyperprolactinaemia with the use of the intervals for prolactin after macroforms separation. J Physiol Pharmacol 2014; 65: 359–364.

37. Samson SL, Hamrahian AH, Ezzat S. American Association of Clinical Endocrinologists. American College of Endocrinology disease state clinical review: clinical relevance of macroprolactin in the absence or presence of true hyperprolactinemia. Endocr Pract 2015; 21: 1427–1435.

