## Supplementary material for "Prevalence of Macroprolactin in Hyperprolactinemic patients receiving Anti-psychotics": Ethical approval

**Sultanate of Oman**
MINISTRY OF HEALTH
Directorate General of Health Services
GOVERNORATE OF MUSCAT
*Director General Office*

سلطنة عُمان
وزارة الصحة
المديرية العامة للخدمات الصحية
لمحافظة مسقط
مكتب المدير العام

Ref: MH/DGHS/DPT/563 / 2020
12 April, 2020

To Dr. Loai Ali

Re/Research and Ethics Committee Feedback

After Compliments,

Reference to your letter regarding your research "Prevalence of macroprolactine in patients receiving anti-psychotics " we are delighted to inform you that the study is approved by the regional research and ethics Committee. Kindly make sure that your data collection won't interfere with the flow of service and remember to send us a copy of the final results.

Your cooperation is highly appreciated

With kind regards

٢٠٢٠/٤/١٩

Dr. Fatma Al Ajmi
Chairperson of Regional Research Committee
Director General of Health Services-Muscat

P.O. Box : 358, Mina Al-Fahal
Postal Code : 116, Sultanate of Oman

ص.ب : ٣٥٨ ميناء الفحل
الرمز البريدي : ١١٦، سلطنة عُمان
فاكس : ٢٤٧٨٢١٠٤ – فاكس : ٢٤٧٨٢١٠٢
